# Feasibility of establishing a serosurveillance system using residual specimens from district-level hospitals: A case study in Choma and Ndola Districts of Zambia

**DOI:** 10.64898/2026.01.08.26343701

**Authors:** Christine Prosperi, Natalya Kostandova, Andrea C. Carcelen, Kalumbu H. Matakala, Innocent C. Bwalya, Gloria Musukwa, Mutinta Hamahuwa, Irene Mutale, Kenny Situtu, Gershom Chongwe, Phillimon Ndubani, Edgar Simulundu, George Chipulu, Joseph Musowoya, Jonathan Kaunda Mwansa, Vickins Malambo, Moses Chakopo, Chaambwa Hachoose, Miriam Kabalo, Emmanuel Luwaya, Auto Madzire, Rebecca Moyo, Clement Chimuka Mukwengo, Tula C. Mwansa, Sandwe Nkoma, Brian Siwakwi, Chipo Syamungulu, Susan Nafukwe, Evans Betha, Lombe Kampamba, Kelvin Kapungu, Japhet Matoba, Passwell Munachoonga, Mathias Muleka, Webster Mufwambi, Sydney Vwalika, Alvira Z. Hasan, Amy K. Winter, Constance Sakala, Francis Dien Mwansa, William J. Moss, Simon Mutembo

## Abstract

Residual blood specimens collected at health care facilities provide a low-cost and readily available specimen source to monitor population immunity through serological surveillance compared to more resource-intensive probability-based surveys. Despite concerns about the representativeness of these specimens, there has recently been increased interest in the use of residual specimens, driven by the need for rapid estimates of seroprevalence during outbreaks to inform response and prevention. Although residual specimen collection is a key component of surveillance systems in some settings, there is limited evidence of its implementation in low-and middle-income countries. We conducted a pilot project at three health facilities in two districts in Zambia between September 2021 through July 2022 to demonstrate the feasibility of residual blood specimen collection integrated into the health system to estimate measles seroprevalence. Through this pilot project, we were able to collect residual pediatric and adult specimens but with some modifications needed if residual specimen collection were to be implemented at scale, such as better integration into the routine functioning of laboratories and health facilities. This paper summarizes our experiences designing and implementing this residual blood specimen collection, including the lessons learned and recommendations for collecting residual specimens from hospitals.

## Introduction

Serological surveillance is the use of blood specimens to test for antibodies such as those that signal exposure or immunity to pathogens of interest. This information can be linked to sociodemographic characteristics and epidemiological data to estimate the burden of disease, identify risk factors for exposure, monitor trends in disease, as well as identify population immunity gaps to vaccine-preventable diseases by age and location (1, 2). The gold standard study design to estimate population-level seroprevalence is a high-quality serological survey that uses probability-based sampling. However, this design requires substantial financial, personnel, transportation, and other resources (2-4). A nationally representative serological survey provides valuable information to develop population immunity estimates at subnational levels, but in many settings they are conducted infrequently (e.g., every four or five years) due to the extensive resource requirements and are specific to a limited number of priority diseases (5-7).

Residual blood specimens collected at health care facilities comprise leftover blood, plasma or serum after clinical testing. These specimens are typically discarded but could provide a low cost, readily available specimen source to monitor population immunity through serological surveillance. The use of residual specimens at health facilities can leverage existing infrastructure because the laboratory equipment and personnel exist and the blood specimens collected for other purposes. Many countries have used residual specimens to monitor seroprevalence to a variety of pathogens, particularly for emerging or reemerging pathogens during an outbreak (8-10). However, there are concerns that epidemiological inferences from residual specimens may be limited because they represent only healthcare-seeking populations (11), and collecting specimens from clinics for patients with specific medical conditions may increase the risk of biases if associated with the disease of interest. Accounting for differences between the patient population attending the health facility and the general population may help improve generalizability.

Residual blood specimens thus provide an alternative or supplementary source of data for serosurveys that are less resource intensive and allow for repeated specimen collection to monitor temporal trends, despite potential limitations in representativeness and the availability of associated sociodemographic data. Considering the tradeoffs between representativeness and data availability and the personnel and financial resource requirements, we envision a multi-pathogen serosurveillance system that leverages diverse specimen sources, including periodic, nationally representative surveys (e.g., Demographic and Health Surveys, Malaria Indicator Surveys, HIV prevalence surveys) and residual specimen collection from health facilities or diagnostic laboratories. We conducted a pilot project in two districts in Zambia to demonstrate the feasibility and utility of integrating residual specimen collection into the health system to estimate measles seroprevalence. We summarize our experience designing and implementing residual specimen collection, including the lessons learned and recommendations for collecting residual specimens at health care facilities.

## Materials and methods

Residual blood specimen collection was conducted in two districts in Zambia: Ndola District, Copperbelt Province and Choma District, Southern Province between 01/09/2021 and 30/06/2022. These two settings were purposively selected based on representing primarily urban and rural areas and to leverage existing partnerships to demonstrate how residual specimens could be collected from health facilities in low- and middle-income countries. We identified tertiary-level facilities drawing patients from across the district where they are located (Table 1): Choma General Hospital in Choma District, and Arthur Davison Children’s Hospital and Ndola Teaching Hospital in Ndola District. District-level hospitals were selected to provide seroprevalence estimates at the district-level. Ethical approvals for residual specimen collection and abstraction of deidentified data were obtained from the Johns Hopkins Bloomberg School of Public Health, the National Health Research and Training Institute (NHRTI), and the Zambia National Health Research Authority. Letters of permission were obtained from the hospitals. No informed consent from patients was required due to abstraction of deidentified data. Lessons learned and recommendations were gathered through direct observations, process mapping, and consultations with key informants involved in collecting residual blood specimens. The measles seroprevalence estimates are presented elsewhere.

**Table 1.** Residual specimen collection, facility, and study characteristics.

|  |  |  |  |
| --- | --- | --- | --- |
|  | Choma General | Arthur Davison<br>Children’s Hospital | Ndola Teaching Hospital |
| District | Choma District | Ndola District |  |
| Facility characteristics (collected during the planning stage) |  |  |  |
| Level of care | Second-level hospital;<br>designated provincial<br>hospital | Third-level hospital;<br>specialty children’s<br>hospital | Third-level hospital |
| Average number of specimens per month received in hematology lab<sup>a</sup> |  |  |  |
| Pediatric | 173 | 1017 | N/A |
| Adult | 1145 | N/A | 4337 |
| Study characteristics |  |  |  |
| Linked institute | Macha Research Trust | National Health Research and Training Institute |  |
| Age groups<br>(years) | 1-4, 5-14, 15+ | 1-4, 5-14 | 15+ |
| Dates of collection | Sept 2021-Jul 2022<sup>a,b</sup> | Oct 2021-May 2022 | Mar-Jun 2022<sup>b</sup> |
| Total number of residual specimens collected |  |  |  |
| 1-4 years | 578 | 530 | N/A |
| 5-14 years | 647 | 612 | N/A |
| 15+ years | 529 | N/A | 431 |

- a. Average number of specimens per month received in hematology laboratory based on inventory of specimens from January, April, July, and October 2019. - b. Average number of specimens per month received in hematology laboratory based on inventory of specimens from January, April, and November 2021.

## Results

We first present the procedures conducted prior to implementation, focusing on how the residual specimen collection steps were adapted for each hospital (Fig 1). We then present details and lessons learned during implementation. Although some examples may be specific to these hospitals, the aim is to provide generalizable lessons learned (Table 2) applicable to different settings.

**Fig 1.**
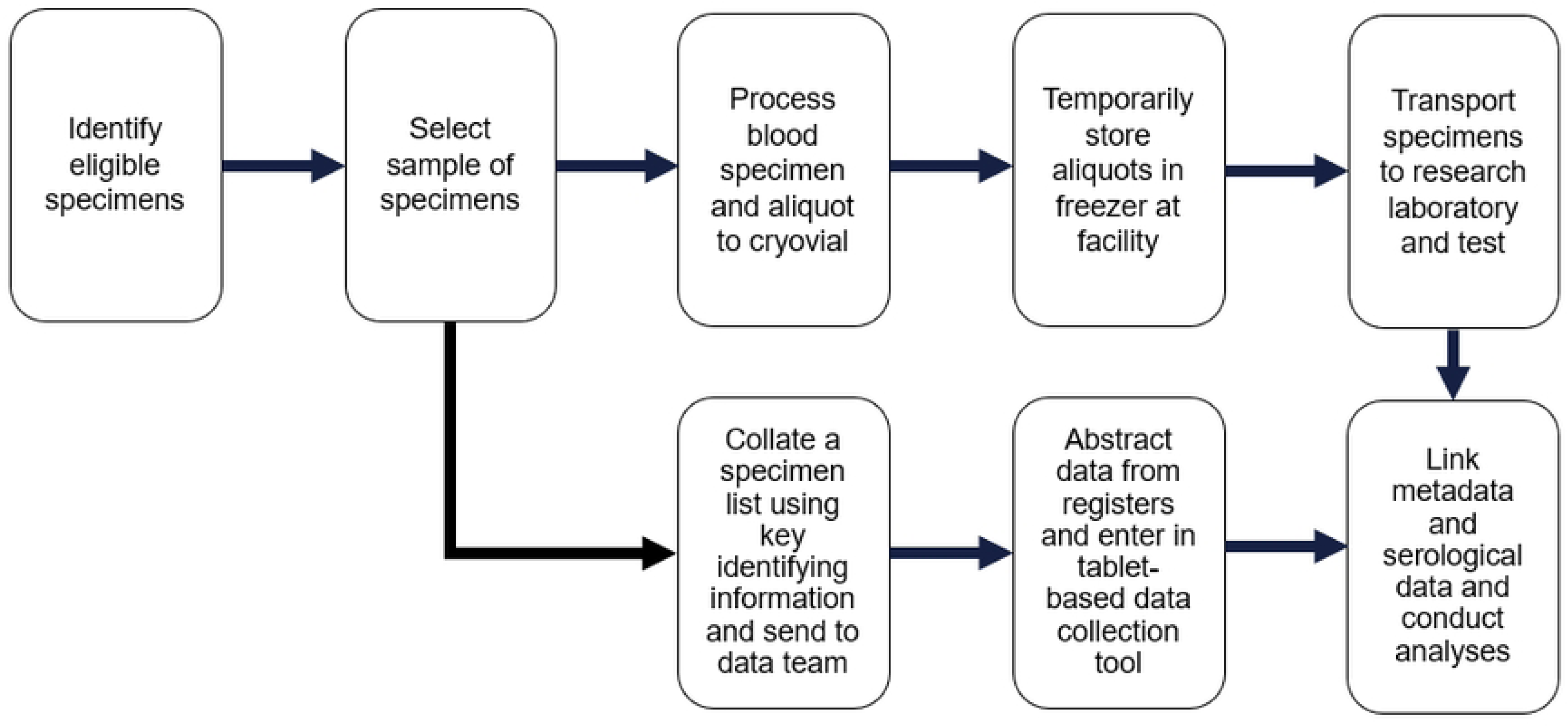
Generic flow of residual specimen collection procedures. Refer to S1 Table for additional details on how procedures were adapted in each facility.

**Table 2.**
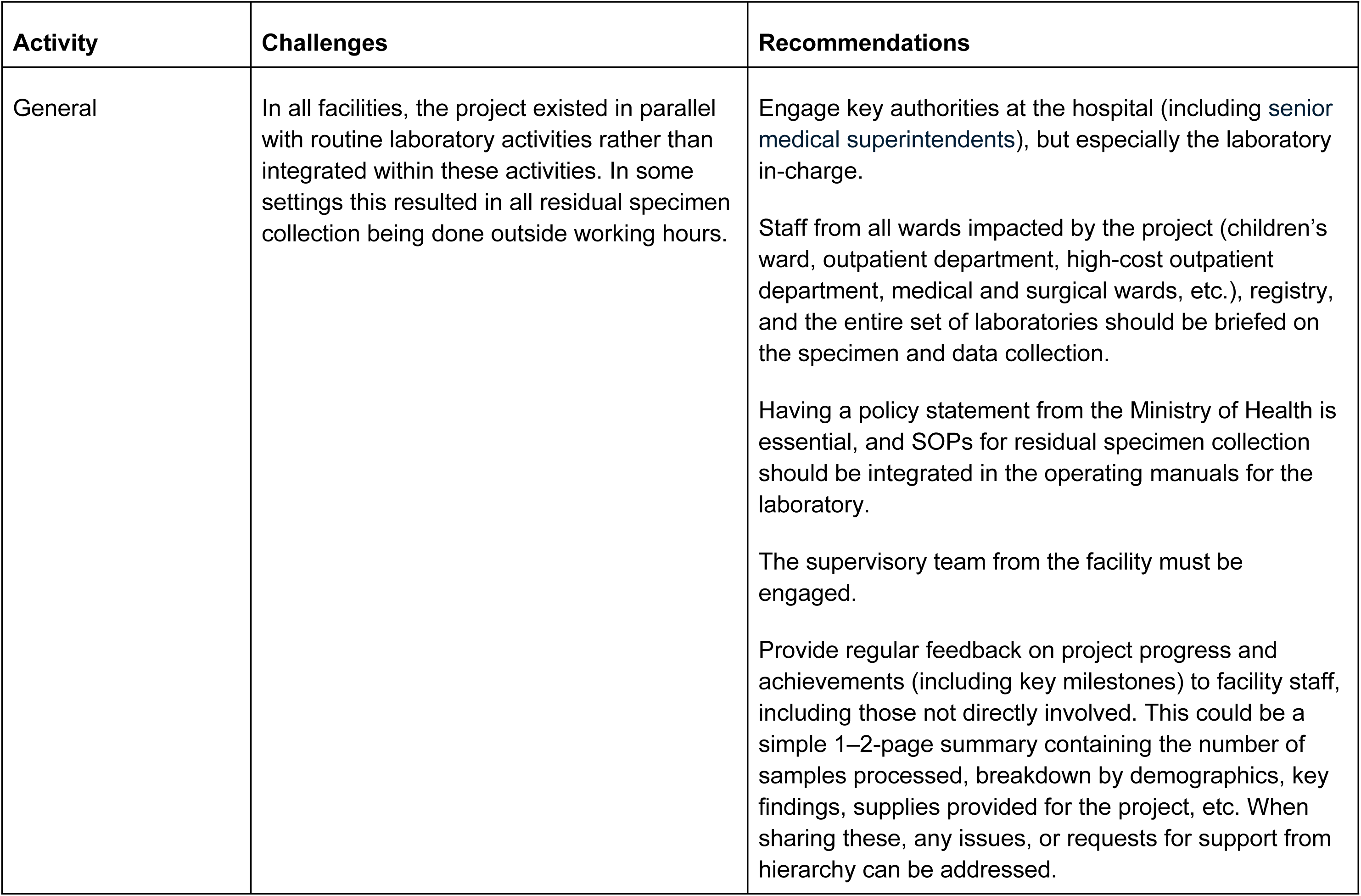
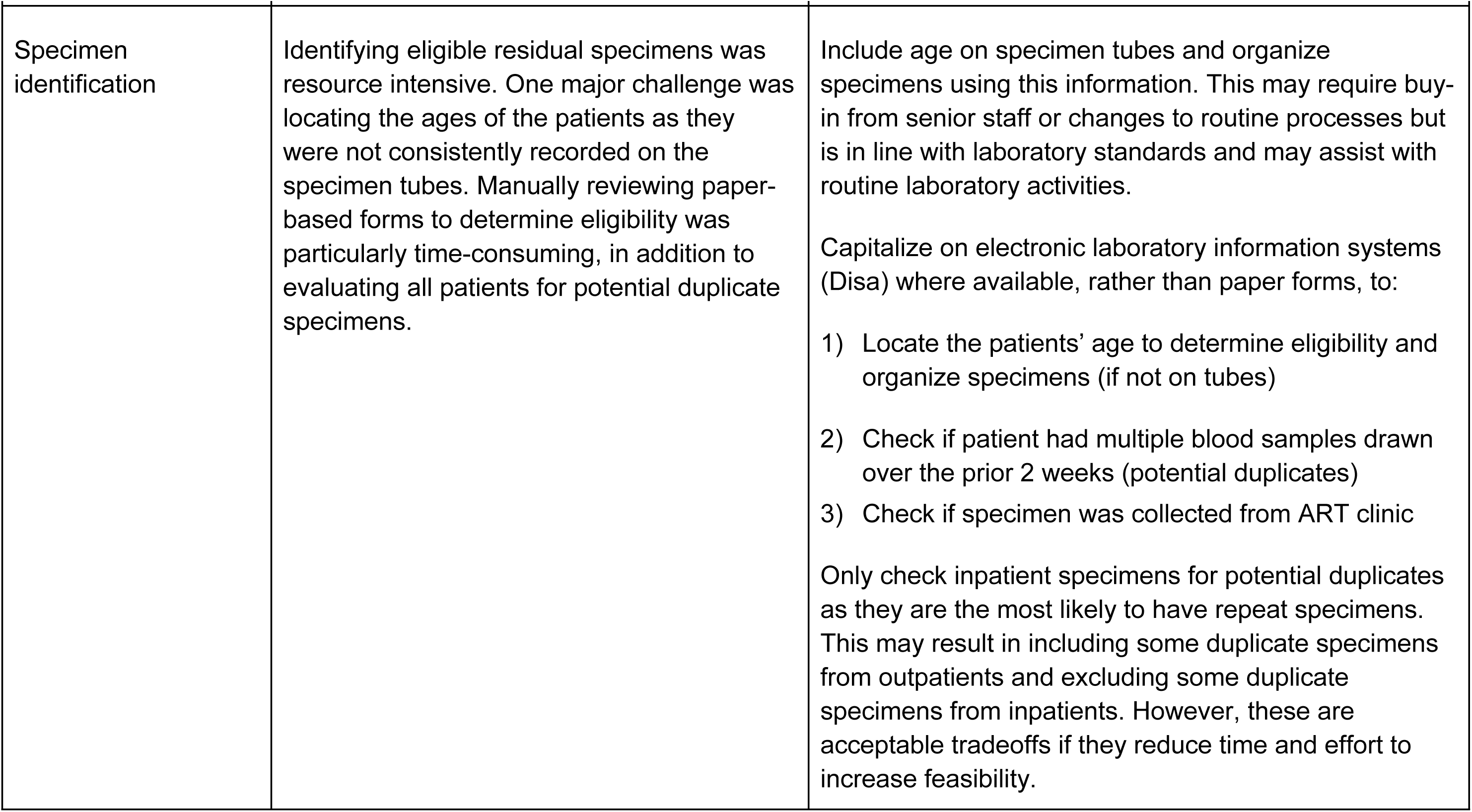
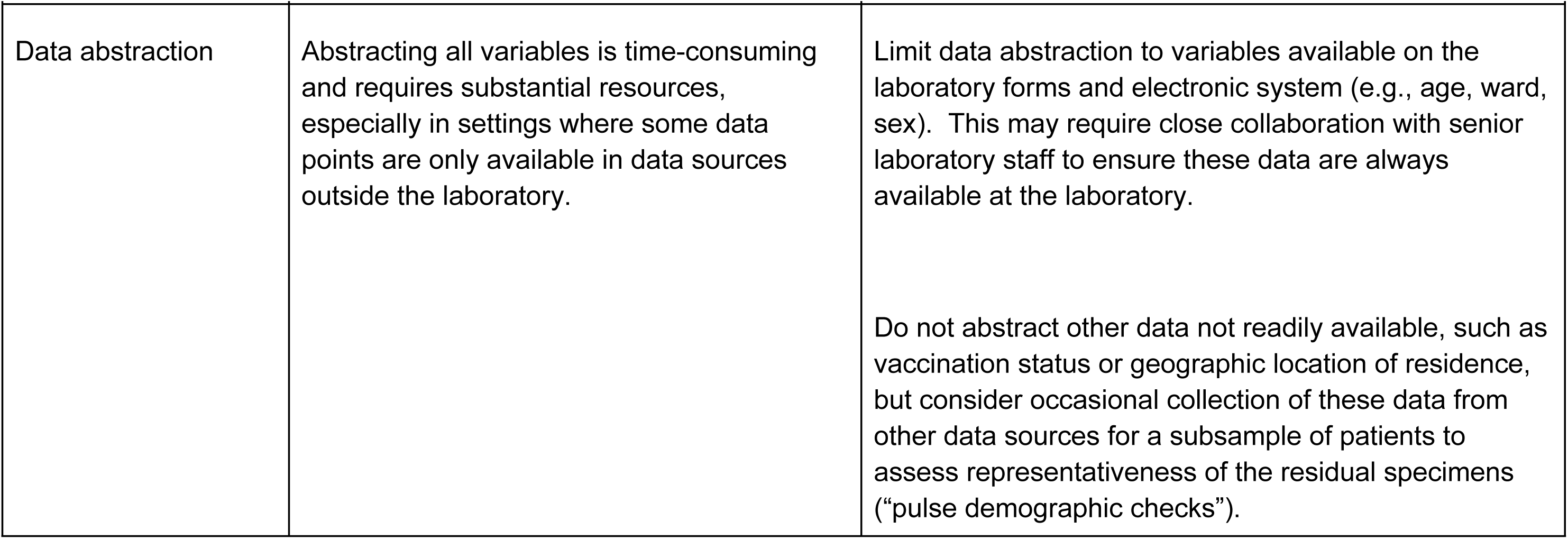
Key lessons learned on implementing facility-based specimen collection.

### Pre-implementation procedures

#### Stakeholder engagement and involvement

The project was implemented by the National Health Research and Training Institute (NHRTI) in Ndola District (formerly Tropical Diseases Research Centre) and Macha Research Trust (MRT) in Choma District. Consultative meetings with hospital staff were held to understand the characteristics and operational details of the hospitals and laboratories to co-create the residual specimen collection that aligns with routine laboratory procedures. These staff included senior management, laboratory management, laboratory technicians, nurses and other clinical staff, and data staff involved in patient registration, data entry, and management of patient files. The main activities during the planning stage were gathering information on patient characteristics, patient referral patterns, patient registration and flow within the hospital, the number of blood specimens collected each month, and blood specimen and data flow. This information was gathered by abstracting aggregated data from facility registers and observing specimen flow at the hospital.

#### Characteristics of patients attending the hospitals

Aggregated characteristics of patients seen at the hospital, such as age, diagnoses, HIV prevalence, and geographic distribution of the health facilities referring the patients, were summarized to understand those seeking care at the hospital and how this population compares to the general population of the district (S2 Table). Data on whether the patient resided within or outside the district or province were limited in the planning stage, so it was not possible to assess if parts of the district were under-represented.

#### Laboratory specimen flow at health facilities

After the hospitals were selected, characteristics of the different laboratories were collected, including the types of whole blood specimen tubes received (e.g., presence of anti-coagulants) and tests performed in each laboratory. Based on this, specialized laboratories, such as microbiology, virology, and parasitology, were excluded. For example, as our primary goal was to estimate measles seroprevalence, we excluded specimens from the virology laboratory which only conducted HIV viral load testing and all specimens received from the antiretroviral treatment (ART) clinic to avoid oversampling specimens from people living with HIV, which could bias seroprevalence estimates. Hematology laboratories were selected as the source of residual specimens given the larger number of blood specimens received and the observation that many specimens sent for biochemistry testing were also sent to the hematology laboratory, potentially resulting in duplicate specimens from the same patient.

#### Specimen characteristics at health facilities

For planning purposes, we compiled the number of specimens received by month, age group, and ward (inpatient or outpatient) for four months during the preceding year, and then estimated the number received per day. In two facilities this information could be summarized using an electronic database but for the other facility this had to be manually summarized using the paper-based register in the laboratory. In laboratories with a large estimated daily number of specimens received, we systematically sampled to reach a daily target and avoid excluding patients based on when their specimens were received in the laboratory. However, in one facility the number of pediatric specimens received in the laboratory each day was small so all eligible specimens were collected.

#### Specimen storage after clinical testing

To develop the protocol, we mapped how the laboratory handled specimens after routine testing was completed, including how the specimens were stored and for how long. Two facilities stored specimens overnight at 2-8°C after testing, so all specimens received in one day were eligible to be processed the following day. One facility stored specimens at room temperature after testing due to limited refrigerator space. To maintain specimen quality at this facility, we restricted specimens to those received on the same day as processing, excluding specimens received between the time of processing and midnight. To assess if these specimens were biased, we purposefully collected “after hours” specimens that would have been excluded because they were collected in the evenings or weekends (depending on the procedure at that facility) during a one-month period for a sensitivity analysis. Additional information was collected during the planning phase on the availability of space and equipment in the laboratory, such as a centrifuge and -20°C freezer, to determine if special arrangements were needed.

#### Data flow at health facilities

To assess the feasibility of data abstraction, we reviewed the laboratory registers and consulted data management staff at the facilities who were familiar with the patient record filing system and data flow to the laboratory. Key data points to be collected for residual specimens included age or date of birth, date of specimen collection, sex, and source of the specimen (inpatient or outpatient), all of which were available in the laboratory registers for most patients. We were also interested in the patient’s place of residence (inside or outside of the district and province; compound or village within the district) and history of measles vaccination for children to explore potential biases in the patient population relative to the district’s general population. However, these variables were not readily available at the laboratory. Separate arrangements, such as engaging the existing health facility data staff and reviewing clinical records, were needed to collect deidentified information for residual specimens. We observed that the availability and source of these data may vary by the type of patient and flow of patients at the facility.

#### Training and initiation

Following the development of procedures based on formative research conducted at each facility, on-site training for laboratory and data management staff was conducted, and supplies for residual specimen collection and processing (e.g., pipette tips, cryovial tubes and boxes, registers) were distributed to the facilities. The training was also used to refine workflow patterns. The local implementing partners remained on site for approximately one week following training to observe procedures and support facility staff.

### Implementation of residual specimen collection at the health facilities

#### Specimen selection and identification of eligible specimens by age

Eligible specimens were defined as whole blood specimens collected in ethylenediaminetetraacetic acid (EDTA)-containing tubes within the age groups of interest (Table 1), collected on the same or prior day and stored at 2-8° C, had sufficient volume (at least 250 μL after centrifugation), and no evidence of severe hemolysis. At each facility, laboratory and data staff were engaged in the residual specimen collection and data abstraction activities, in addition to their routine responsibilities (e.g., processing specimens, performing routine clinical and diagnostic testing, registering patients, preparing surveillance reports).

Although the generic flow of residual specimens was similar across facilities (Fig 1), the specific procedures for identifying specimens were influenced by how specimens were stored after testing, the daily number of specimens received in the laboratory, age groups of interest, and the format of specimen metadata in the laboratory (S1 Table). For example, one facility organized specimen tubes by age group to facilitate systematic sampling, which required abstracting age from the electronic laboratory information system as age was not available on the specimen tubes. A second facility primarily relied on paper-based laboratory request forms to identify eligible specimens. Where age was required to organize specimens, either from electronic or paper-based forms, identifying age-eligible individuals and locating or selecting specimens required at least one hour. In the facility where only adult specimens were selected, specimens were first systematically sampled using the specimen tubes, then age eligibility was confirmed using the electronic record system, which reduced the time required for specimen identification by about half.

Recording the patient’s age on the specimen tube labels would enable technicians to easily organize specimens and substantially reduce the time to identify and select eligible specimens. However, this would require considerable support and buy-in from facility management to implement as a standard practice or to improve adherence if already implemented but not consistently done. Alternatively, specimens could be organized by age groups as routine testing is completed so they are properly arranged for residual processing, or an electronic record system could be used.

#### Quality control checks for specimens

Once specimens were selected, they were manually cross-checked against the list of specimens pulled during the prior two weeks to determine if the patient’s sample was recently processed as a residual specimen; if so, the specimen was excluded to reduce biases in the seroprevalence estimate. A two-week window was selected to reduce burden when searching for duplicates. Specimens were also checked to ensure eligibility criteria were met. Excluded specimens were replaced if additional eligible specimens were available.

Most duplicates were from inpatients. Limiting duplicates check to inpatient specimens could improve efficiency. Using an electronic medical record system to assess if the selected patient had multiple specimens sent to the laboratory within the past two weeks would further streamline the process of excluding duplicate specimens.

#### Specimen Processing

The selected whole blood specimens were labeled with study identification numbers, covering any identifiable information on the tube, centrifuged, aliquoted into two labeled cryovials, placed in freezer boxes, and stored at -80° C until transported to the central laboratory for testing.

The frequency of specimen transport from the health facility to the laboratory at the research institutes differed based on the distance between the two locations. For facilities located far from the laboratory, specimen transport was combined with trips for other purposes.

### Data abstraction

Laboratory staff provided the list of patients whose specimens were collected that day to data staff to abstract data. The procedures for data abstraction were generally influenced by how data were recorded and stored at the hospital, which varied by type of patient and data point. The abstracted data were entered into a tablet-based application (KoboToolBox) by either the laboratory team or the data team. In some situations, the data were first abstracted onto a paper register by the data team and then shared with the laboratory team for data entry.

#### Data systems

There were many challenges identified in the data abstraction process. Data staff needed to access multiple data sources to abstract the requested data for a single specimen. One hospital had an electronic laboratory management system but locating and abstracting linked data for residual specimens using this system was challenging. Therefore, staff needed to manually search records to abstract data from the medical registers or in the individual patient files. At a second hospital, an electronic medical record system was available for outpatients but data for all inpatients and certain data points for outpatients needed to be abstracted from the paper-based ward registers. At the third hospital, all data abstraction was done using the paper-based ward registers, except for the patient’s referring facility. This information was only available at registration; however, where registration took place varied based on how the patient entered the hospital. In some situations, data abstraction staff needed to go to other sections located far apart to access the relevant registers.

Nurses did not report disruptions due to the presence of the data staff manually searching records; however, moving records within the facility could be problematic. The staff made multiple attempts to locate records. For example, if the record had not been filed or was with the medical staff during the first attempt, staff needed to adapt to the timing of ward rounds.

Complex filing systems for patients using private insurance also complicated the process of locating records for individuals insured under someone else’s plan. The staff encountered missing records because some patients took their records and others entered the hospital through private arrangements and were not properly registered. On average, locating the patient files and data points took 1 to 3 days including reattempts, but at times required up to a week.

Staff reported that the process of locating records became more challenging as more time passed since collection, so timely communication between the laboratory and data teams was important.

Modifying procedures for recordkeeping, such as requiring clinicians to provide prescription notes so patients do not need to take their medical records to the pharmacy may benefit both clinical care and residual specimen data abstraction.

#### Data points

Certain data points were more problematic to abstract than others (S3 Table). Measles vaccination status proved especially challenging, requiring staff to access individual patient files and separate registers. At one facility, few children had vaccination data available because it was not consistently recorded by clinicians. The other facility reported high under-5 card availability for pediatric patients, although clinicians only recorded if the child was fully immunized or up to date; in these circumstances, we assumed the child had received the measles vaccine. Missing data on age were more common for adults where it may simply be recorded as “adult” if the patient did not know their date of birth and was unable to estimate their age. Missing data were also more common for patients in the emergency department, especially if the person was not accompanied by a close relative who knew that information.

Limiting the requested data to those recorded on the laboratory request form would greatly simplify data abstraction, substantially reduce the time required to collect key information, and facilitate implementation of a residual specimen system that is more sustainable. All data abstraction could be completed in the laboratory by the laboratory staff, removing the requirement for a separate data team or the need to access multiple data sources and minimize the time staff had access to identifiable information. In facilities where age is not consistently recorded on laboratory request forms, this would require significant collaboration with the laboratory manager and facility leadership to reinforce that this information must be recorded and to reject specimens missing this information. If there are concerns about the representativeness of the seroprevalence results, periodic “pulse demographic data checks” could be conducted to abstract data on variables not available at the laboratory, such as referring facility, place of residence (e.g., district, village or ward), and vaccination status. These data could be used to explore changes over time in the patient characteristics, which may influence interpretation of the seroprevalence estimates obtained from the residual specimens.

#### Data staff and resources

Utilizing data staff familiar with the systems at the facility was critical to facilitate data abstraction, especially given the complexity of locating and accessing registers and clinical records. Having sufficient data staff was important to ensure staff were available to support data abstraction without disrupting routine patient registration.

Other logistical challenges included access to reliable internet to access the electronic medical record system and upload data. Lack of dedicated computers for the study meant that staff had to wait for the workstations to become available to abstract data from the electronic medical record system. Providing dedicated computers and internet bundles to support data abstraction would facilitate data abstraction.

### Ethical considerations

The residual specimens were de-identified and relabeled with a study identification number at the time of processing. However, the link between the study identification number and patient identifiers was temporarily maintained on paper-based logs securely stored at the health facility to facilitate locating the patient records and abstracting linked metadata. There was no interaction with the patients, and no identifiable information was collected from patient records. Although the date of birth was initially abstracted, it was used to calculate age in years, after which the birth date field was removed from the database.

## Discussion

This project demonstrated the feasibility of residual blood specimen collection to conduct serosurveillance for vaccine-preventable and other diseases at hospitals in Ndola and Choma Districts in Zambia without disrupting the operations of the laboratory or hospital information system. We highlighted key aspects of the process, from planning to specimen selection to abstraction of metadata from patient laboratory and medical records. We also identified challenges as well as proposed solutions for future residual specimen collection at hospitals. Some of these solutions require health system operational changes or investments in technological advancements that could provide benefits to the hospitals beyond supporting residual specimen collection. Other minor suggestions could facilitate the implementation of residual specimen collection at other hospitals.

This pilot study was conducted at hospitals with minimum standards of a level 2 or higher to leverage laboratory equipment, trained personnel, and health information systems that may be more efficient than those at lower-level health centers or clinics. The ideal facility for residual specimen surveillance should have characteristics such as an established and fully functioning laboratory that can store whole blood for 1-2 days, refrigerator or freezer space for the temporary storage of specimens at the appropriate temperature, electronic laboratory management system, functioning equipment such as a centrifuge, reliable electricity or back-up generator, and hardwired internet. Anticipated costs for residual specimen collection include supplies for residual specimens and processing, transport to the central laboratory, management personnel, serological testing supplies, and laboratory personnel (S4 Table). In many settings, the facilities meeting these characteristics may be limited to larger, district- or regional-level hospitals. However, adaptations to procedures could be made to facilitate collection at other facilities to allow for a broader representation of the population.

The timing and frequency of residual collection may be determined by the goals of the system and diseases of interest, in addition to logistical considerations. Unlike other surveillance systems designed to trigger specimen collection for individuals meeting specific criteria (e.g., suspected measles cases or individuals with influenza-like illness), residual specimens from selected health facilities or diagnostic laboratories can be continually collected from the population seeking care at these facilities. The selected health facilities thus act as sentinel sitesfor ongoing residual specimen collection and testing for emerging and re-emerging pathogens, ideally in a setting where the collection is fully integrated with routine laboratory activities (12).

Unlike research designed to purposefully collect specimens requiring informed consent from participants, the ethical considerations for secondary use of residual specimens are ambiguous (10). Patients seeking care at facilities may not be aware or intentionally consent to additional testing of their specimens. Anonymizing specimens can mitigate the confidentiality risks to participants but means that the results cannot be provided back to the individuals. We tested for historical exposure to vaccine preventable diseases, specifically measles, SARS-CoV-2, and tetanus. For most individuals tested their individual results would not influence future behaviors or actions, but younger children could have the opportunity for catch-up vaccination. If testing were expanded to include other diseases where actions could be taken to prevent spread or mitigate disease, concerns could arise that the results would need to be reported back to participants. This would mean specimens could not be anonymized if results needed to be provided to participants (13).

For sustainability at the hospital level, residual specimen collection should be integrated with routine laboratory activities rather than in parallel. This ensures laboratory staff can complete activities during their standard workday, prevents disruptions to ongoing activities, and creates opportunities to adapt routine procedures to integrate residual specimen collection. Engaging all staff at the hospital in the residual specimen collection, including those not directly involved in specimen collection such as clinical, registry, and staff in other labs at the health facility, would frame the work as surveillance rather than a research project. This also provides opportunities for staff indirectly involved to provide feedback on how the program may be affecting their work so problems can be identified and addressed promptly. Training a broader set of staff also ensures the ability to transition tasks in the event of staff turnover. Regular feedback from the external management teams at the regional level or public health institutes to the hospital about progress and key findings will help facilitate continued engagement of the staff and create platforms to discuss issues with supplies, procedures, or additional support required by the facility.

In addition, residual specimen collection should be viewed as a formal part of the nationwide surveillance system similar to those for other infectious diseases (e.g., fever-rash for measles and rubella; HIV; influenza), with policy guidelines from the Ministry of Health on collection, storage, and ethical considerations of using residual specimens for the purpose of disease surveillance. Procedures and systems, including oversight and transport logistics, should be adapted from existing surveillance systems or fully integrated, where possible (14). For example, the hub and spoke model used by other surveillance systems in Zambia could be used to transport residual specimens from peripheral facilities to a district facility and then to a central laboratory (15). Discussions with program managers during the planning process should also include how seroprevalence estimates could be used to complement the Integrated Disease Surveillance and Response system to guide program and policy. The use of residual specimens will become increasingly important given funding constraints that may limit valuable resources that have been leveraged in the past for serosurveillance, such as Demographic Health Surveys and Demographic Surveillance Systems (16).

Residual blood specimens collected at health facilities as part of a serosurveillance system could provide valuable epidemiologic information to help guide public health programs and policies. There are key data points and information that are helpful for planning the residual specimen collection process, which may differ based on existing health facility procedures. During the design phase, consideration should be made to balance feasibility of residual specimen collection with the value of the information generated. Building on the feasibility and lessons learned in Zambia, other countries should consider residual specimen collection from health facilities as an additional surveillance platform.

## Data Availability

The underlying data are qualitative in nature and are only available upon request to the corresponding author if requested within 3 years of the publication date.

## Acknowledgments

We greatly appreciate the support from all health facility staff who assisted in the planning and implementation of residual specimen collection at the selected facilities, including Danny Chilambwe (Records Medical Officer, Choma General Hospital), Chilulwe Kasonde (PCR molecular laboratory in-charge, Choma General).

## Supporting information

**S1 Table. Variability in residual specimen procedures by facility S2 Table. Characteristics collected during planning stage**

**S3 Table. Data abstracted from health records**

a. Data variables that were more difficult to obtain because they required going to a different part of the hospital.

b. Date of birth was only used to calculate age and was not retained in the data system.

**S4 Table. Resource considerations for planning residual biospecimen collection**

